# A DISSYMMETRY IN THE FIGURES RELATED TO THE COVID-19 PANDEMIC IN THE WORLD: WHAT FACTORS EXPLAIN THE DIFFERENCE BETWEEN AFRICA AND THE REST OF THE WORLD?

**DOI:** 10.1101/2020.05.17.20104687

**Authors:** Cheikh Faye, Cheikh Tidiane Wade, Ibrahima Demba Dione

## Abstract

Humanity has experienced outbreaks for millennia, from epidemics limited to pandemics that have claimed many victims and changed the course of civilizations. The advent of vaccines has eradicated some of the serious pathogens and reduced many others. However, pandemics are still part of our modern world, as we continue to have pandemics as devastating as HIV and as alarming as severe acute respiratory syndrome, Ebola and the Middle East respiratory syndrome. The Covid-19 epidemic with 0-exponential contamination curves reaching 3 million confirmed cases should not have come as a surprise, nor should it have been the last pandemic in the world. In this article, we try to summarize the lost opportunities as well as the lessons learned, hoping that we can do better in the future. The objective of this study is to relate the situation of Covid-19 in African countries with those of the countries most affected by the pandemic. It also allows us to verify how, according to the observed situation, the African ecosystem seems to be much more resilient compared to that of other continents where the number of deaths is in the thousands. To verify this, the diagnosed morbidity and mortality reported for different states of the world are compared to the ages of life and the average annual temperature of these states. The results show that the less dramatic balance of the African continent compared to other continents is partly linked to the relatively high temperatures on the continent but also to the relatively young character of its population.

## 1. Introduction

Humanity has experienced outbreaks for millennia, from epidemics limited to universal pandemics that have claimed many victims and changed the course of civilizations. The advent of vaccines has eradicated some of the serious human pathogens and mitigated many others. However, pandemics are still part of our modern world, as we continue to have pandemics as devastating as HIV and as alarming as Severe Acute Respiratory Syndrome, Ebola and Middle East Respiratory Syndrome (**Saqr and Wasson, 2020**). The Covid-19 epidemic with exponential curves reaching 3 million confirmed cases should not have come as a surprise. However, we seemed to ignore the past. (**Peeri *et al*., 2020; Morse, 2007**). Unfortunately, Covid-19 is not the last pandemic in the world and we need to learn what we missed and how to avoid failures.

The world is more globalized and more vulnerable: connectivity has “dissolved” the boundaries between countries transcending the barriers of distance. While the disease is believed to have started in one city in China, it has spread to all continents, despite administrative boundaries. The situation is very different from the 1918 influenza pandemic known as the Spanish flu, where travel and urbanization were much less pronounced than today. While border closures and travel restrictions within countries may be helpful, this is much less effective than in the past. Pandemics require a stronger WHO with sufficient resources (**Morse, 2007**). Failure to manage a pandemic in one country can have repercussions for the entire planet; therefore, pandemics require more solidarity and coordination so that fragile countries can find the resources to treat, isolate and combat severe epidemics. There are good signs that such efforts are being implemented (for example, the European Union has announced EUR 15 billion to combat the current pandemic in developing countries) and, hopefully, these efforts are being consolidated to become systemic, proactive and organized. In other words, pandemics require global efforts with a strong and resourceful World Health Organisation.

Having appeared in China in the City of Wuhan, Covid-19, initially a zoonosis, has spread throughout most of the world to become a pandemic affecting all social strata and relatively all ages of life. Today, more than 3 million people are affected and the spread of Covid-19 continues to grow beyond the world’s best performing health systems. However, it is clear that the geography of Covid-19 disease shows significant disparities between countries and age groups in terms of the level of disease and the extent of mortality. This differentiated prevalence prompts reflection on possible explanations by taking into account a set of endogenous and exogenous factors (geographical, environmental, biological, socio-cultural, political contexts, etc.).

New epidemiological trends on transmission and mortality in Africa and the most affected regions of the world suggest that better studies of this infection in sub-Saharan Africa than in other regions of the world are needed. The COVID-19 pandemic has lower rates of local transmission and mortality in Africa, the region where the virus was the last to arrive (**Imaralu, 2020**). The daily statistics emanating from the high infectious property of the new strain of coronavirus Covid-19, particularly its rapid worldwide transmission and the nature of the resulting deaths sweeping across countries, call for concerted efforts to limit local transmission in already colonized territories. There is currently no known consensual cure for COVID-19 infection and there is currently no evidence to recommend specific anti-Covid-19 treatment for patients confirmed to have this disease (**WHO, 2020a**).

The news of very high mortality rates in industrialized countries with stronger health systems and sophisticated infrastructure is cause for concern (**WHO, 2020b**). Facilities and equipment in industrialized countries that have so far provided assistance to developing countries are overwhelmed and not even sufficient to meet the current challenges facing these countries. As of 13 April 2020, 1,850,527 confirmed cases of Covid-19-positive persons and 114,245 deaths worldwide had been reported to WHO (**WHO, 2020c**). There was concern about the impact of this virus on African nations, given the weakness of prevailing health systems with suboptimal infrastructural support (**WHO, 2020c**). The recent mass exodus of health workers to Europe and the Americas and the continuing security threats of terrorism and violent crime make this new deadly viral threat a source of concern for governments in this subregion (**Imaralu, 2020**).

Recent trends in transmission, morbidity and mortality worldwide, however, seem to be at odds with the predictions of misery for the people of sub-Saharan Africa. Current statistics that place Africa in the least affected region (as of 13 April 2020) with 14525 cases and 788 deaths raise many questions given the particular social and infrastructural vulnerability of this region (**WHO, 2020c**). The elderly, people with other co-morbid chronic diseases such as lung, kidney, heart disease and diabetes mellitus, especially diseases affecting ACE-2 signalling, are at increased risk of Covid-19 infection and adverse outcomes (**Huang *et al*., 2020**).

On the basis of the above, the social and health implications of intervention measures to limit the spread of Covid-19 virus should be considered and interventions carefully planned000. This study thus proposes a diachronic reading of the evolution of Covid-19 with as inputs the diagnosed morbidity and mortality reported in different States of the world on the one hand, and on the other hand the life expectancy and the average annual temperature of these States for a comparative study in order to draw all the specificities generated. This contribution focuses on the factors explaining this disparity in a statistical, sociodemographic and geographical analysis. It is based on a statistical treatment of aggregated data with a plural input taking into account the specificities of the prevalence of Covid-19 at the level of the countries of the world.

## 2. Materials and methods

### 2.1. Data

The Covid-19 statistics used in this study are from the World Health Organization database and are as of Monday, April 13, 2020 (https://www.weather-atlas.com or http://data.un.org/Data.aspx?d=CLINO=ElementCode%3A11). The temperature data are from the Climate Research Unit, located at the University of East Anglia in the United Kingdom (https://fr.wikipedia.org/wiki/Liste_des_pays_par_temp%C3%A9rature_moyenne). Population structure data are from the World Bank database (https://donnees.banquemondiale.org/pays/senegal?view=chart), as of 2019 (Table 1 and Figure 1).

**Figure 1:**
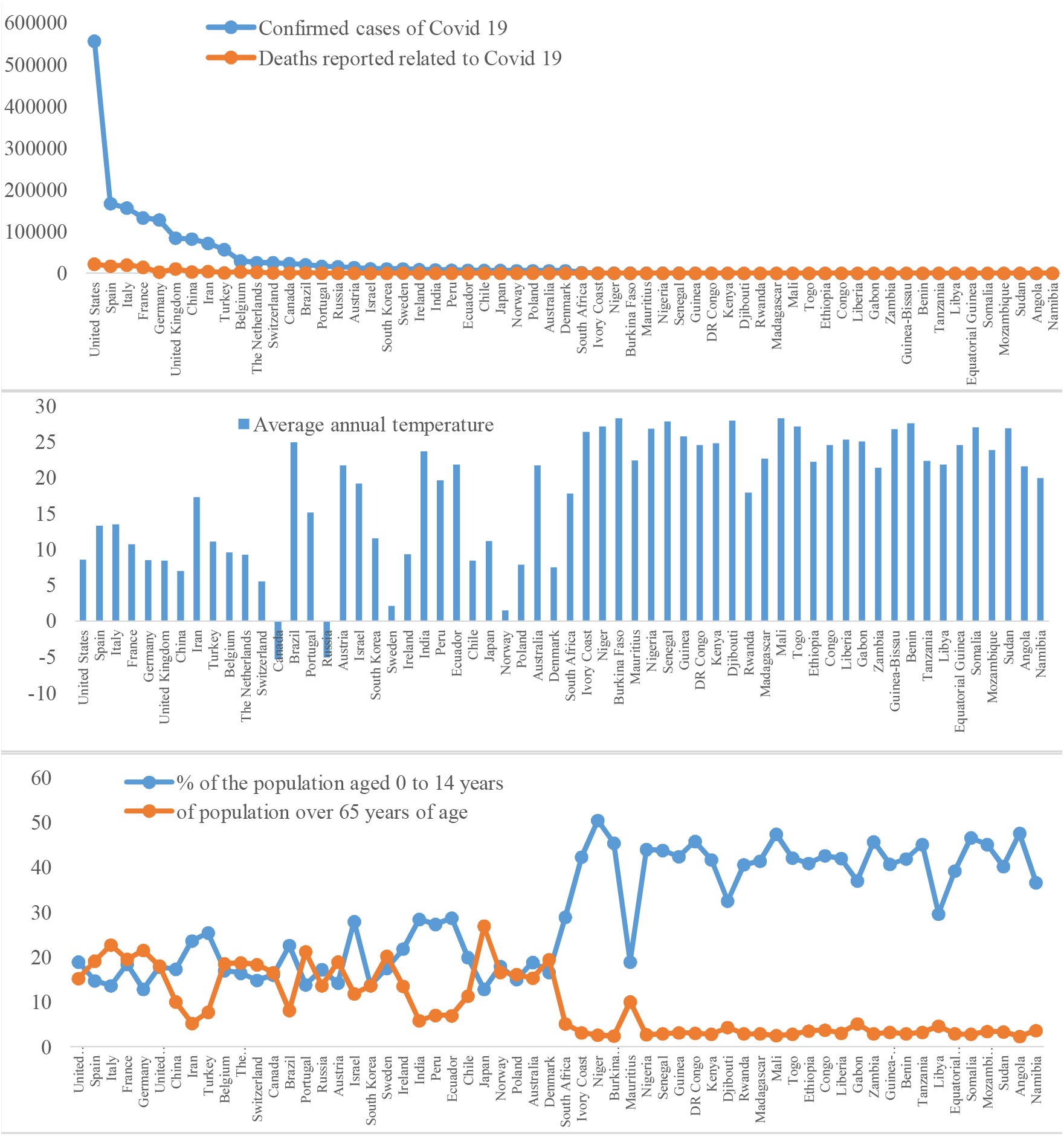
Covid-19, temperature and population structure data for the countries selected for this study

For this study, 60 countries were selected, 30 countries in Africa (these are indeed the African countries that have counted the most cases of Covid-19 as of Monday 13 April 2020) and 30 countries generally located in other continents (18 countries in Europe, 6 in America, 5 in Asia, 1 in Oceania). Indeed, these are the 30 countries in the world that have counted the most cases of Covid-19 as of Monday 13 April 2020. The objective of this study is to relate the situation of Covid-19 in African countries with those of the countries most affected by the pandemic. It also allows us to verify in what way the African ecosystem seems, according to the observed situation, much more resistant compared to that of other continents where the number of deaths is counted in thousands. To verify this state of affairs, two key hypotheses are raised: one natural (this is the average annual temperature of the country) and the other anthropogenic (this is the structure of the population). According to several scientists, the less dramatic balance of the African continent compared to other continents would be linked to the relatively high temperatures on the continent but also to the relatively young character of its population. To carry out this correlation study, the data used are shown in the following table.

**Table 1:** Covid-19, temperature and population structure data for the countries selected for this study

| Selected countries |  | Covid-19 situation of Monday 13 April 2020 |  | Climate framework | Structure of the country's population |  |  |
| --- | --- | --- | --- | --- | --- | --- | --- |
| Country | Country No. | Confirmed cases | Declared Deaths | Temperature | From 0 to 14 years old | From 15 to 64 years old | Over 65 years old |
| United States | 1 | 555398 | 22023 | 8,55 | 18,9 | 65,9 | 15,2 |
| Spain | 2 | 166831 | 17209 | 13,3 | 14,7 | 66,2 | 19,1 |
| Italy | 3 | 156363 | 19899 | 13,45 | 13,6 | 63,7 | 22,7 |
| France | 4 | 132591 | 14393 | 10,7 | 18,4 | 62,1 | 19,5 |
| Germany | 5 | 127854 | 3022 | 8,5 | 12,8 | 65,7 | 21,5 |
| United Kingdom | 6 | 84279 | 10612 | 8,45 | 17,8 | 64,2 | 18 |
| China | 7 | 82160 | 3341 | 6,95 | 17,3 | 72,7 | 10 |
| Iran | 8 | 71686 | 4474 | 17,25 | 23,6 | 71,2 | 5,2 |
| Turkey | **9** | 56956 | 1198 | 11,1 | 25,4 | 66,9 | 7,7 |
| Belgium | **10** | 29647 | 3600 | 9,55 | 17 | 64,5 | 18,5 |
| The Netherlands | **11** | 25587 | 2737 | 9,25 | 16,4 | 64,9 | 18,7 |
| Switzerland | **12** | 25300 | 987 | 5,5 | 14,8 | 66,9 | 18,3 |
| Canada | **13** | 23318 | 653 | -5,35 | 16 | 67,5 | 16,5 |
| Brazil | **14** | 20727 | 1124 | 24,95 | 22,6 | 69,3 | 8,1 |
| Portugal | **15** | 16585 | 504 | 15,15 | 13,8 | 65 | 21,2 |
| Russia | **16** | 15770 | 130 | -5,1 | 17,2 | 69,2 | 13,6 |
| Austria | **17** | 13945 | 350 | 21,65 | 14,2 | 66,9 | 18,9 |
| Israel | **18** | 10878 | 103 | 19,2 | 27,9 | 60,6 | 11,8 |
| South Korea | **19** | 10537 | 217 | 11,5 | 13,7 | 72,7 | 13,6 |
| Sweden | **20** | 10483 | 899 | 2,1 | 17,5 | 62,3 | 20,2 |
| Ireland | **21** | 8928 | 320 | 9,3 | 21,8 | 64,7 | 13,5 |
| India | **22** | 8447 | 273 | 23,65 | 28,4 | 65,8 | 5,8 |
| Peru | **23** | 7519 | 193 | 19,6 | 27,3 | 65,4 | 7 |
| Ecuador | **24** | 7257 | 315 | 21,85 | 28,7 | 64,4 | 6,9 |
| Chile | **25** | 6927 | 73 | 8,45 | 19,9 | 68,8 | 11,3 |
| Japan | **26** | 6926 | 132 | 11,15 | 12,8 | 60,3 | 26,9 |
| Norway | **27** | 6415 | 119 | 1,5 | 17,9 | 65,5 | 16,6 |
| Poland | **28** | 6356 | 208 | 7,85 | 15 | 68,9 | 16,1 |
| Australia | **29** | 6322 | 61 | 21,65 | 18,8 | 65,9 | 15,3 |
| Denmark | **30** | 6174 | 273 | 7,5 | 16,6 | 64 | 19,4 |
| South Africa | **31** | 2028 | 25 | 17,75 | 28,9 | 66 | 5,1 |
| Côte d'Ivoire | **32** | 533 | 4 | 26,35 | 42,3 | 54,6 | 3,1 |
| Niger | **33** | 491 | 11 | 27,15 | 50,5 | 45,9 | 2,6 |
| Burkina Faso | **34** | 484 | 27 | 28,25 | 45,4 | 52,2 | 2,4 |
| Mauritius | **35** | 319 | 9 | 22,4 | 18,9 | 71,1 | 10 |
| Nigeria | **36** | 318 | 10 | 26,8 | 44 | 53,3 | 2,7 |
| Senegal | **37** | 280 | 2 | 27,85 | 43,8 | 53,3 | 2,9 |
| Guinea | **38** | 250 | 0 | 25,7 | 42,4 | 54,5 | 3,1 |
| DR Congo | **39** | 234 | 20 | 24,55 | 45,8 | 51,2 | 3 |
| Kenya | **40** | 197 | 8 | 24,75 | 41,7 | 55,5 | 2,8 |
| Djibouti | **41** | 187 | 2 | 28 | 32,5 | 63,3 | 4,3 |
| Rwanda | **42** | 120 | 0 | 17,85 | 40,6 | 56,5 | 2,9 |
| Madagascar | **43** | 106 | 0 | 22,65 | 41,4 | 55,7 | 2,9 |
| Mali | **44** | 105 | 9 | 28,25 | 47,4 | 50 | 2,5 |
| Togo | **45** | 76 | 3 | 27,15 | 42,1 | 55,1 | 2,8 |
| Ethiopia | **46** | 71 | 3 | 22,2 | 40,9 | 55,6 | 3,5 |
| Congo | **47** | 60 | 7 | 24,55 | 42,6 | 53,7 | 3,7 |
| Liberia | **48** | 50 | 5 | 25,3 | 42 | 55 | 3 |
| Gabon | **49** | 49 | 1 | 25,05 | 37 | 57,9 | 5,1 |
| Zambia | **50** | 40 | 2 | 21,4 | 45,7 | 51,4 | 2,9 |
| Guinea-Bissau | **51** | 38 | 0 | 26,75 | 40,7 | 56,1 | 3,2 |
| Benin | **52** | 35 | 1 | 27,55 | 41,9 | 55,2 | 2,9 |
| Tanzania | **53** | 32 | 3 | 22,35 | 45,1 | 51,7 | 3,2 |
| Libya | **54** | 25 | 1 | 21,8 | 29,6 | 65,8 | 4,6 |
| Equatorial Guinea | **55** | 21 | 0 | 24,55 | 39,2 | 57,9 | 2,9 |
| Somalia | **56** | 21 | 1 | 27,05 | 46,6 | 50,6 | 2,8 |
| Mozambique | **57** | 21 | 0 | 23,8 | 45,1 | 51,5 | 3,4 |
| Sudan | 58 | 19 | 2 | 26,9 | 40,2 | 56,5 | 3,3 |
| Angola | 59 | 19 | 2 | 21,55 | 47,6 | 50,1 | 2,3 |
| Namibia | 60 | 16 | 0 | 19,95 | 36,6 | 59,8 | 3,6 |

### 2.2 Methods

Principal component analysis (PCA) is a widely used statistical technique (**Pulido-bosch et al. 1999; Helena et al. 1999; Tidjani et al. 2006; Eslamian et al. 2010; Faye 2014; Baba-Hamed and Bouanan 2016**). It reduces the number of variables to those that are the most significant among a set of variables and is used to find a link between variables and individuals in order to group them into homogeneous regions. One of the objectives of PCA is to obtain useful information from a data matrix, and to provide a graphical representation of the data to facilitate analysis. The mathematical procedure of principal component analysis is actually a multivariate statistical method of data processing.

We subjected all the variables studied for the different countries under study to a principal components analysis, in order to determine the affinities between these countries and to deduce the most characteristic parameters. To do this, a correlation matrix was used and the components were determined according to the type of rotation of the orthogonal axes. The first factorial axis (F1) of this representation is such that it determines the maximum inertia of the cloud and thus the variance. The second axis (F2) perpendicular to the first expresses the maximum remaining variance. The third axis, always perpendicular to the other two, is defined by the maximum remaining inertia; etc.

Principal Component Analysis, or PCA, is a method of reducing the number of variables allowing the geometric representation of observations and variables. This reduction is only possible if the initial variables are not independent and have non-zero correlation coefficients (Bouroche and Saporta, 1980). The method was applied to 60 countries (30 in Africa and 30 in other continents) and 6 variables which are: the Covid-19 situation on Monday 13 April 2020 (confirmed cases and reported deaths), the mean annual temperature and the structure of the proportion (0 to 14 years, 15 to 64 years and over 65 years).

## 3. Results

The final reconstitution of the distribution of the countries allowed us to define the factor axes or factors responsible for this distribution and consequently, to highlight the affinities between the different countries and to deduce the variables linked to the Covid-19 pandemic that best characterize them.

### 3.1 Matrix of correlation coefficients

Analysis of Tables 2 and 3 and the eigenvalue curve (Figure. 2), shows that the first three factors represent the maximum amount of information. Thus the first three factor axes express 93.3% of the total variance, with 63.17% for the first factor, 23.24% for the second and 6.8% for the third factor (Table 2 and Figure 2).

**Table 2:** Eigenvalues of the correlation matrix between Covid-19, temperature and population structure data of the countries selected for this study

|  | Own value | total variance | cumulative % |
| --- | --- | --- | --- |
| Axis 1 | 3,79 | 63,17 | 63,17 |
| Axis 2 | 1,39 | 23,24 | 86,42 |
| Axis 3 | 0,41 | 6,87 | 93,30 |

**Table 3:** Correlation matrix between Covid-19, temperature and population structure data for the countries selected for this study

| Variables | Confirmed cases | Declared Deaths | Temperature | From 0 to 14 years old | From 15 to 64 years old | Over 65 years old |
| --- | --- | --- | --- | --- | --- | --- |
| Confirmed cases | 1 |  |  |  |  |  |
| Declared Deaths | 0,85 | 1 |  |  |  |  |
| Temperature | -0,31 | -0,30 | 1 |  |  |  |
| From 0 to 14 years old | -0,35 | -0,39 | 0,79 | 1 |  |  |
| From 15 to 64 years old | 0,27 | 0,25 | -0,65 | -0,89 | 1 |  |
| Over 65 years old | 0,36 | 0,45 | -0,76 | -0,91 | 0,6 | 1 |

**Figure 2:**
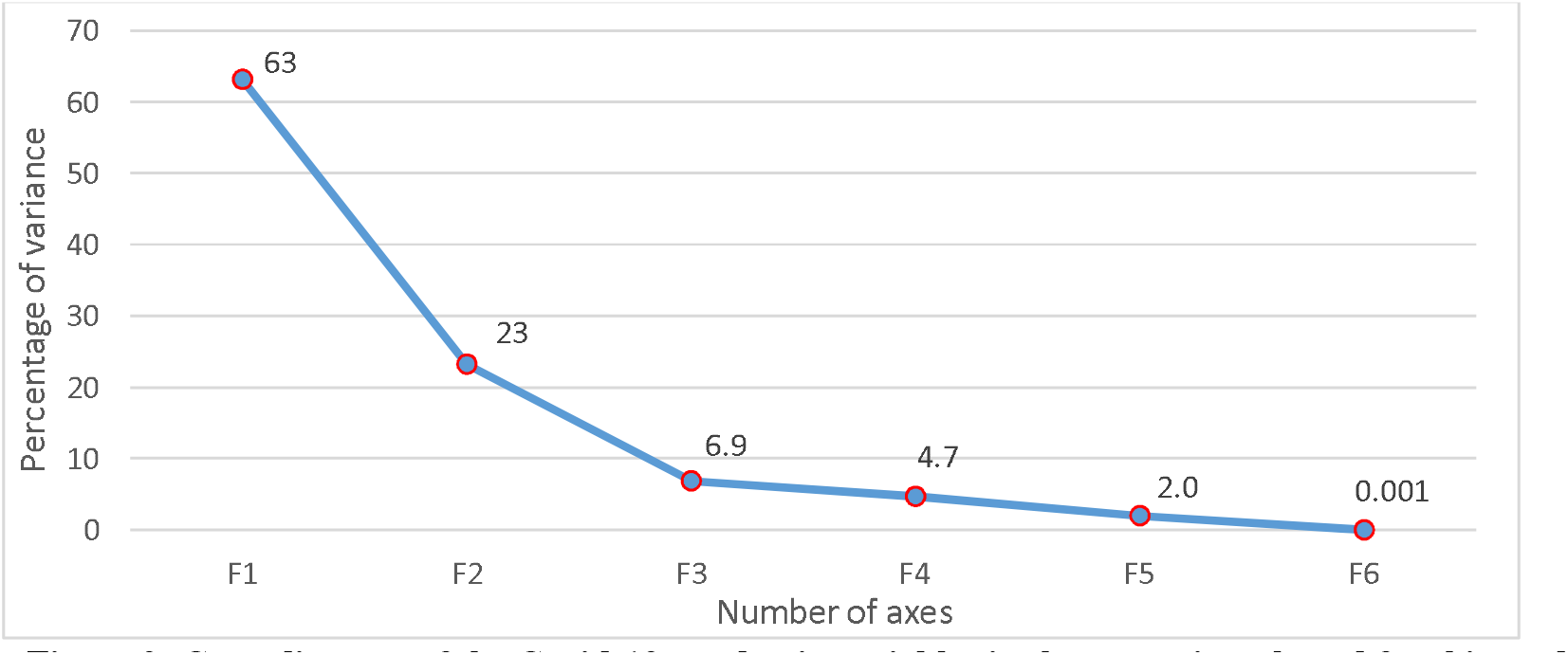
Cone diagram of the Covid-19 pandemic variables in the countries selected for this study

Analysis of the correlation matrix of Covid-19 pandemic variables shows that:

There is an inverse relationship between the Covid-19 pandemic (confirmed cases and reported deaths) and temperature, suggesting that generally the warmer a country is, the less likely it is to be affected by the Coronavirus pandemic, and vice versa.

The opposing relationship between the Covid-19 pandemic (confirmed cases and reported deaths) and the proportion of elderly (and/or young) is apparent, suggesting that generally the younger a country’s population is (and therefore the fewer vulnerable people), the less likely it is to be affected by the Coronavirus pandemic, and vice versa.

The number of confirmed cases is well correlated with the number of deaths, weakly correlated with the ageing character of the population and negatively correlated with the average temperature of the country and the youthful character of the countries’ population.

The axes 1, 2 and 3 thus selected highlight their relations with the parameters studied (Table 4).

**Table 4:** Vectors of the correlation matrix of selected Covid-19 pandemic variables at the country level

| Variables | F1 | F2 | F3 |
| --- | --- | --- | --- |
| Confirmed cases | -0,31 | 0,63 | 0,16 |
| Declared Deaths | -0,32 | 0,62 | -0,04 |
| Temperature | 0,43 | 0,23 | 0,43 |
| From 0 to 14 years old | 0,49 | 0,23 | -0,12 |
| From 15 to 64 years old | -0,42 | -0,29 | 0,74 |
| Over 65 years old | -0,46 | -0,14 | -0,48 |

The correlation matrix of the Covid-19 pandemic variables and variance weight factors (Table 4) and Figure. 3 show that axis 1 (main axis of inertia) which represents more than 63.17% variance is very well positively correlated with the mean annual temperature of the country and the proportion of young people in the total population of the country. It is also weakly but negatively correlated with the number of confirmed Covid-19 cases, the number of reported deaths and the proportion of elderly in the total population.

**Figure 3:**
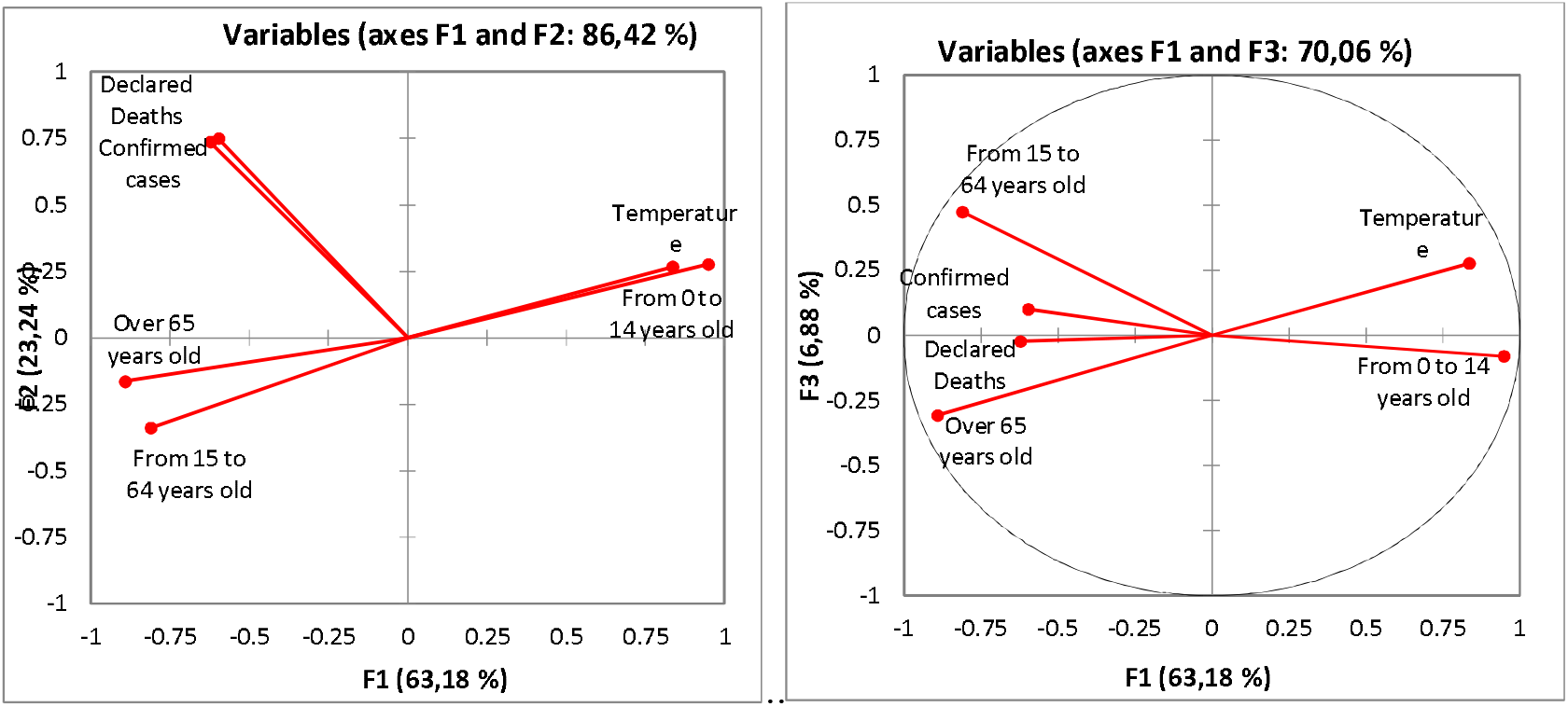
Diagram of the contributions of selected Covid-19 pandemic-related variables at the country level for factor 1, 2 and 3

Axis 2, with 23.24% variance, is positively related, strongly with the number of confirmed Covid-19 cases, the number of reported deaths and the proportion of elderly in the total population, weakly with the average annual temperature of the country and the proportion of young people in the total population of the country. It is negatively correlated, albeit weakly, with the proportion of the elderly in the total population.

As for Axis 3, which represents almost 6.8% of the variance, it shows only a positive but weak correlation with the number of confirmed Covid-19 cases and the mean annual temperature of the country. Its correlation with the number of reported deaths and the proportion of young and old in the total population remains negative. The correlation of the Covid-19 pandemic variables studied with this axis, whether positive or negative, is weak.

These different characteristics of the Covid-19 pandemic-related variables and of the countries studied are represented in circles and planes, respectively (Figures 3 and 4), which illustrate the projection of the variables and the variables on factorial planes 1 and 2.

**Figure 4:**
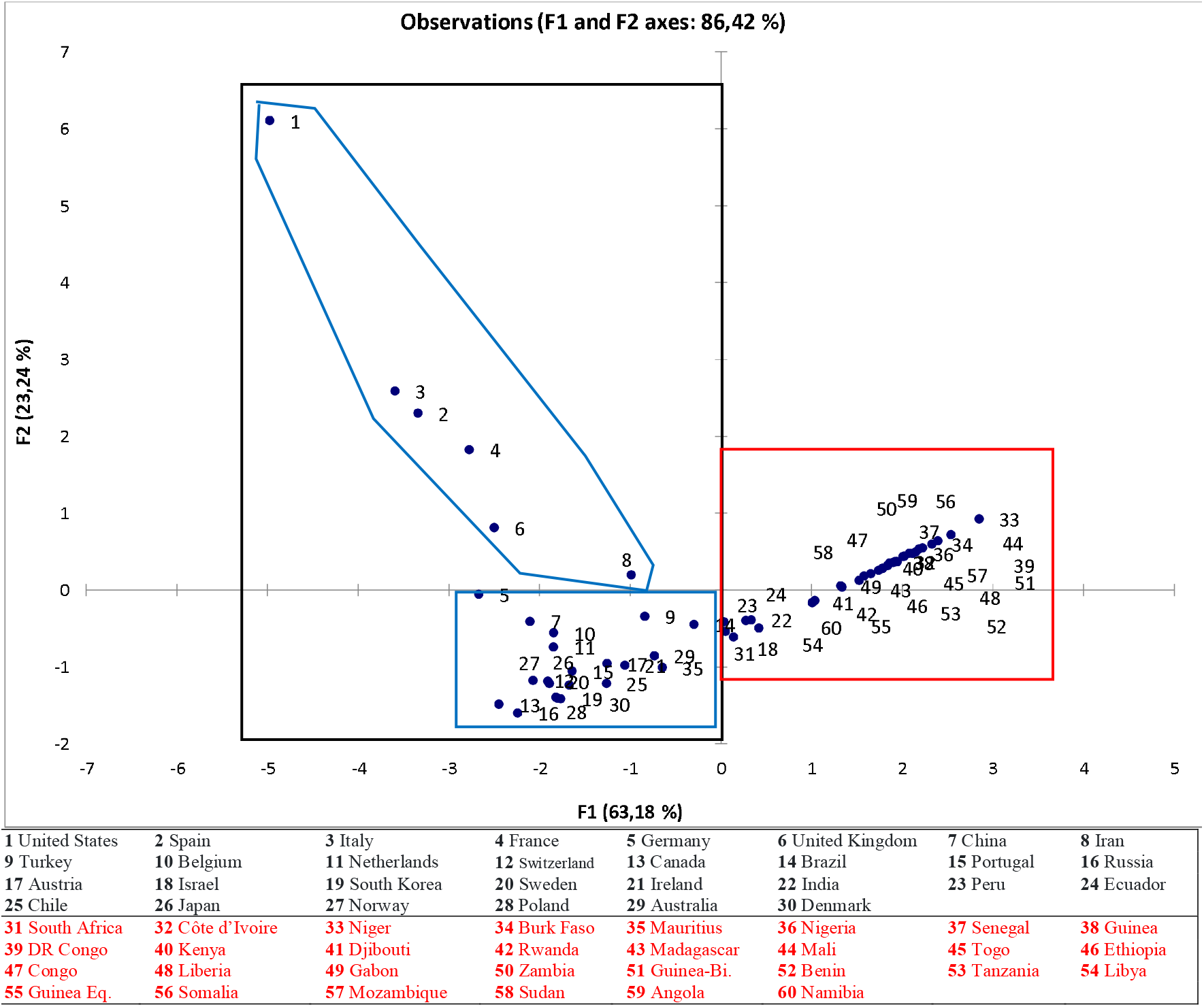
Projections of selected countries on factor 1 and 2 plans

### 3.2 Characteristics of the variables according to the three axes

Each variable related to the Covid-19 pandemic is associated with a point whose coordinate on a factorial axis is a measure of the correlation between that variable and the factor (axis 1 or axis 2 or axis 3). Projected on a factorial plane, the variables studied fit into a side plane 2 (Figure 3). They are all the closer to the side of the plane the more the variable is well represented by the factorial plane, i.e. the variable is well correlated with the two factors making up this plane.

In Plans I-II and I-III (63.18% of cumulative inertia), **Axis I** is determined by variables related to the Covid-19 pandemic such as the mean annual temperature of the country and the proportion of young people in the total population, which are opposed to the number of confirmed Covid-19 cases, the number of reported deaths and the proportion of elderly people in the total population.

### 3.3 Country characteristics by axis

The purpose of this representation is to provide approximate planar images of the cloud of the countries located in the plane. Thus, the x-axis represents the thermal component (mean annual temperature of the country) and the anthropogenic component (proportion of young people in the total population) of the countries, while the y-axis represents their profile (in terms of number of confirmed Covid-19 cases and number of reported deaths). Indeed, a country represented on the positive part of axis 1 generally has a high temperature and a young character of its population; this is the case, for example, of African countries.

In the U.S. plan, Plan I-II highlights three groups. The first group is made up of the countries that have recorded the greatest number of cases of contamination and deaths (e.g. United States, Spain, Italy, Germany, France, etc.). These are the countries with the highest form values (A, P, kg, L and l). In this group, the United States stands out clearly from the others due to its higher number of contamination and deaths. The second group is made up of European countries (Belgium, Netherlands, Switzerland…), American countries (Canada, Brazil, Peru…), Asian countries (Israel, South Korea, Japan…) and Oceania (Australia) which have recorded a lower number of cases of contamination and deaths than those in the first group, despite certain common characteristics (such as temperature and the ageing of the population). The last group is made up of African countries with the lowest number of cases and deaths related to Covid-19. These countries also benefit from a high temperature and a young population.

What analysis can be made of this CPA Figure 4, which distinguishes three categories of countries?

- The first category (usually 1-8) and the second category (usually 9-31) are made up of the European, American and Asian countries most affected by the Covid-19 pandemic. These countries are generally located in the temperate zone (where temperatures are relatively mild and conducive to the easy spread of the pandemic). In addition, they are countries with an aging population, and therefore a high number of vulnerable people), hence the high number of deaths related to the pandemic. However, there are some exceptions because among these countries there are a few that are located in the tropical zone, such as Brazil, and are in this group. Some African countries, such as Mauritius, are also located in the group, due to its low average annual temperature, which brings it closer to countries in the temperate zone.
- The third category (generally 31-60) consists strictly of African countries selected for this study (those most affected by the Covid-19 pandemic). These countries are generally located in the tropical zone (whose temperatures are relatively high and unfavourable to the spread of the pandemic). In the early stages of the epidemic, the speed of contagion decreases with the temperature of the country or region and high temperatures reduce the initial rates of contagion (**Demongeot *et al*., 2020**). Containment policies and other rules of expulsion should take into account climatic heterogeneities in order to adapt public health decisions to possible geographical or seasonal gradients. In addition, these are countries with young populations (i.e., with very few vulnerable people), hence the low number of pandemic-related deaths.

Ultimately, the Principal Component Analysis (PCA) synthesized the information contained in Table 1 by crossing countries (individuals) and variables (quantitative). It produced a summary of information (Figures 1, 2, 3 and 4) by establishing similarity between the selected countries, searching for homogeneous groups of countries, identifying a typology of countries and variables related to the Covid-19 pandemic studied, and also identifying linkage budgets between variables related to the Covid-19 pandemic, using synthetic parameters or variables. The CPA has generally established the linkages between these two typologies (**Kouani *et al*., 2007**).

In conclusion, we note that the CPA has the advantage, on the one hand, of summarizing the set of correlated initial parameters into a reduced number of uncorrelated factors. On the other hand, it has enabled us to highlight similarities or oppositions between parameters and sub-basins **(Faye, 2014; Baba-Hamed and Bouanan, 2016**) What analysis can be made of this CPA Figure 4, which distinguishes three categories of countries?

- The first category (usually 1-8) and the second category (usually 9-31) are made up of the European, American and Asian countries most affected by the Covid-19 pandemic. These countries are generally located in the temperate zone (where temperatures are relatively mild and conducive to the easy spread of the pandemic). In addition, they are countries with an aging population, and therefore a high number of vulnerable people), hence the high number of deaths related to the pandemic. However, there are some exceptions because among these countries there are a few that are located in the tropical zone, such as Brazil, and are in this group. Some African countries, such as Mauritius, are also located in the group, due to its low average annual temperature, which brings it closer to countries in the temperate zone.
- The third category (generally 31-60) consists strictly of African countries selected for this study (those most affected by the Covid-19 pandemic). These countries are generally located in the tropical zone (whose temperatures are relatively high and unfavourable to the spread of the pandemic). In the early stages of the epidemic, the speed of contagion decreases with the temperature of the country or region and high temperatures reduce the initial rates of contagion (**Demongeot *et al*., 2020**). Containment policies and other rules of expulsion should take into account climatic heterogeneities in order to adapt public health decisions to possible geographical or seasonal gradients. In addition, these are countries with young populations (i.e., with very few vulnerable people), hence the low number of pandemic-related deaths.

Ultimately, the Principal Component Analysis (PCA) synthesized the information contained in Table 1 by crossing countries (individuals) and variables (quantitative). It produced a summary of information (Figures 1, 2, 3 and 4) by establishing similarity between the selected countries, searching for homogeneous groups of countries, identifying a typology of countries and variables related to the Covid-19 pandemic studied, and also identifying linkage budgets between variables related to the Covid-19 pandemic, using synthetic parameters or variables. The CPA has generally established the linkages between these two typologies (**Kouani *et al*., 2007**).

In conclusion, we note that the CPA has the advantage, on the one hand, of summarizing the set of correlated initial parameters into a reduced number of uncorrelated factors. On the other hand, it has enabled us to highlight similarities or oppositions between parameters and sub-basins **(Faye, 2014; Baba-Hamed and Bouanan, 2016**).

## 4. Discussions

Two months after the first cases of Covid-19 appeared in Africa, the spread of the disease appears to be progressing more slowly than elsewhere. Since the first cases of Covid-19 infection detected in Africa in mid-February, as of ^1^ May 2020, there have been just over 26,663 reported cases (including those already cured) and 973 deaths in Africa, compared with over 3,175,207 cases of illness and 224,172 deaths worldwide (**WHO, 2020d)**. Statistically, many experts still point to the African anomaly and link it to climate, geography and, in the most extreme cases, even to a kind of biological resilience (**Savana, 2020**). Africa, with 17 per cent of the world’s population, is home to only 0.83 per cent of the world’s sick and 0.7 per cent of its dead. Better still, with already more than 12,000 recoveries, it seems to be much more resistant than others to Covid-19. For the time being, in any case, no one denies that the spread of the virus seems to be singularly slow on the continent, and many reasons are cited to try to explain this (**Marbot, 2020)**.

**Climate:** Like influenza, coronavirus is believed to be a disease that thrives in the cold season and does not tolerate heat, drought, or even heavy sun exposure. The hypothesis seems to be supported by the fact that the countries most affected by the pandemic have a rather temperate climate and that most cases are concentrated either in the extreme north of the continent or in the extreme south, where heat and drought are less overwhelming. On the research side, a British study confirms that, on average, fewer respiratory illnesses are found in hot and dry countries, and an American report of April 24 states that the half-life of the virus, i.e., the period required for its infectious power to be halved, may increase from 18 to 6 hours if heat and humidity increase (**Marbot, 2020)00**. Researchers nevertheless remain very cautious, like the director of international affairs at the Pasteur Institute, Pierre-Marie Girard, who stresses that during in vitro experiments it was found that the coronavirus “multiplied very well in heat”. Sun, heat and humidity could weaken the Covid-19 virus. According to a study of the American government, presented Thursday April 23 in Washington, the virus responsible for the pandemic of Covid-19 weakens in a hot and humid atmosphere as well as under the rays of the sun. *“Our most striking observation to date is the powerful effect that sunlight seems to have in killing the virus, both on surfaces and in the air,”* said a senior Department of Homeland Security official **Bill Bryan (2020)**. Despite this, Health Minister Olivier Véran was sceptical and the WHO believes that high temperatures do not prevent the virus from being contracted.

**The youth of the population:** This is the other major explanation put forward. In English-speaking countries, it has even become a slogan: *“The virus isold and cold and Africa is young and hot”*. Doctors confirm that the majority of severe cases of Covid-19 involve people over 60 years of age, which would be fortunate for the continent, where the median age is 19.4 years and 60% of the population is under 25 years of age. It is also pointed out that one of the hardest hit countries, Italy, has 23.1 per cent of its population aged 65 and over, compared to 5 per cent in Africa. There is almost unanimous agreement on this hypothesis, but scientists qualify it by pointing out that although the African population is young, it is unfortunately more affected than others by diseases such as HIV or malnutrition, which can make it vulnerable. Finally, some researchers note that in Europe and the United States the elderly often live among themselves in old people’s homes, which encourages the spread, whereas in Africa they more frequently live with their families. This could protect them.

**Less dense habitat: With the** exception of a few countries such as South Africa, Egypt, Morocco or Algeria, and some large megacities, population density is on average lower in Africa than in the parts of the world where the coronavirus has wreaked the greatest havoc (Western Europe and North America). On average, there are 42.5 inhabitants per km2 in Africa, compared to 207 in Italy and… more than 10,000 in New York State. The WHO confirms that this is a positive factor, while pointing out that these figures are only an average, and that cities such as Lagos or Abuja have record population densities. Today, this position must be put into perspective because in some countries, it can be observed that most cases of infection concern localities with dense populations, including cities.

**More limited displacement:** Another rational explanation that is difficult to circumvent is that the African population moves less, on average, than that of many advanced countries, and the risks of contamination are therefore necessarily lower. As a reminder, there is only one African airport in the list of the world’s 50 sites with the highest concentration of air traffic: Johannesburg.

**The experience of epidemics: As** many point out, this is not the first epidemic in Africa, and there have been far more deadly ones, including the recent Ebola crises. Healthcare workers and populations alike are therefore used to dealing with health crises, lessons have been learned and “good practices” have been put in place. Certain methods of detection, isolation of patients, and precautions during care developed previously are duplicable in the face of the coronavirus. The authorities also took the measure of the danger more quickly than others and put in place very early on the control or closure of borders, distancing or containment. This led Dr Moumouni Kinda, who has faced several crises with the non-governmental organization ALIMA, to say, “Epidemics like Ebola have given us experience on communication and awareness, which are key points in breaking the chains of transmission of the virus.

**True cross-border cooperation:** For some African scientists, the continent also has the advantage of practicing true solidarity. When one country lacks masks or test kits, neighbouring countries that are less affected are likely to provide them. Lesotho, which does not yet have an operational laboratory, has its samples tested in South Africa, and a network for detecting seasonal influenza, used against Covid-19, already brings together some 20 countries on the continent. Without being overly optimistic, it must be said that solidarity sometimes seems to work better in Africa than in certain richer regions, where we see the major laboratories jealously watching over their discoveries in the hope of being able to market a treatment or a vaccine. Not to mention a Donald Trump trying to get his hands on the patents of drugs under development for the sole (financial) benefit of the United States…On a much more local scale, it is also pointed out that the community-based functioning of many African populations makes it possible to better convey prevention messages, but also to detect patients more quickly, since few people are likely to be left to their own devices.

**Indirect protection from other treatments:** This hypothesis is the subject of much controversy, and the WHO, in particular, is very cautious. However, some doctors have noted some disturbing coincidences: there are reportedly fewer coronavirus contaminations in the countries most affected by malaria” or tuberculosis. Or in those that massively vaccinate their population with BCG. Would contracting certain diseases be a barrier to Covid-19? It will take time to prove it, but many doctors believe that antimalarial treatments such as chloroquine have some effectiveness. That’s partly why French Professor Didier Raoult and teams like the Drug Discovery and Development Centre (H3D) at the University of Cape Town are giving priority to testing antimalarial drugs. The WHO is critical, noting that some countries such as Burkina Faso, Nigeria and Senegal, where malaria is devastating, are not spared by the virus. More recently (20 April), the Malagasy President announced that his country was in possession of a *“vita malagasy”* remedy (*made in Madagascar*) *with* preventive and curative virtues against the coronavirus. Covid-Organics, the name given to this treatment, is an herbal tea made from dried artemisia leaves, produced by the Malagasy Institute for Applied Research (IMRA). Despite WHO warnings, member countries of the Economic Community of West African States (ECOWAS) will now be able to treat their coronavirus patients with Covid-Organics, and test the effectiveness of this improved traditional remedy proposed by IMRA. Faced with the fight against the Covid-19 pandemic, Africans have chosen to put forward the unity and solidarity of Africa. Madagascar was able to demonstrate to the world that we Africans can cooperate and help each other not only in an economic situation but above all in a health and humanitarian situation.

**A “genetic” immunity:** What if Africans were protected by their DNA, which, for some reason yet to be determined, would be more robust against the coronavirus? The hypothesis is far from unanimous - at the Pasteur Institute, Pierre-Marie Girard “doesn’t really see why” such a specificity would exist - and will take time to be explored. The Cameroonian professor Christian Happi, specialist in genomics, who divides his time between Harvard University and Nigeria, does not completely rule out this possibility: “Africans are exposed to many diseases, so it is possible that their bodies react better. You’ll have to look for antibodies to find out, but it’s possible. After Ebola, we saw that many people in Nigeria had been exposed to the disease but had not developed it. »

**Another version of the virus: An** idea that is similar to the previous one: since it now seems that several different strains of Covid-9 are present on the planet (up to eight distinct forms), perhaps the one present in Africa is less aggressive? This could also explain the fact that there seem to be more asymptomatic cases there than elsewhere. The hypothesis remains audacious insofar as the virus arrived through patients who contracted it elsewhere. Could it have mutated afterwards? The WHO does not rule out the idea, but stresses that in order to validate it it will be necessary to sequence the genome of Covid-19, which is currently underway.

**Better masks:** When asked about the specificities that could work in favour of Africa, MatshidisoMoeti, WHO Africa Director, points out that the continent “has a very active and competent textile industry”, especially in Brazzaville, where the organization’s offices are located. This particularity perhaps allows the population to benefit from more and better quality cloth masks than in some rich countries, where scarcity is the rule.

In conclusion, the scientists point out that what probably explains the low number of cases observed on the continent is, above all, the fact that most countries took drastic protection measures very early on. And perhaps also the fact that because the disease initially affected people who were travelling, rather better informed than average and living mostly in cities, it was easier to identify the first cases than in other epidemics. But the modesty of the figures continues to amaze, as Congolese biologist Francine Ntoumi notes: *“In some countries on the continent, people eat bats, people live on top of each other… In fact, everything is done to make it explode, but it doesn* t. “It’s up to African scientists to find out why.

However, caution should be exercised in the face of the figures because even if Africa is not the continent most affected by Covid-19, the damage could be considerable, according to virologist Denis Chopera. In addition, the contamination rate would be underestimated due to the lack of medical facilities. In the context of this SARS-COV-2 virus, transmission can take place during the last days of viral incubation, before symptoms appear or at least are significant. This is a viral strategy that has certainly allowed the virus to spread so impressively. Another risk of under-evaluating cases is the lack of diagnostic tests or one of the reagents that make them up. Given the international demand, the whole world is struggling to obtain everything necessary to fight the pandemic and Africa is not always in an ideal position to be able to negotiate prices compared to other regions of the world.

At the moment, patient management is going relatively well in the hospitals that have been identified to play this role and the system is not overwhelmed, although some tools, such as respirators, are sorely lacking. At present (as of May ^1^ 2020), just under 1000 (973) deaths for approximately 27 times the number of identified confirmed cases, which would correspond to a lethality of 3.6%. Although the global case-fatality rate is generally higher (7%), severe cases are more easily identified than non-symptomatic or minimally symptomatic cases, and it is possible that several foci of the infection have not been identified. If this is the case and the virus is insidiously transmitted in the population, it is likely that hospital infrastructures will be rapidly overwhelmed when the weakest are affected.

## 5. Conclusion

Globally, deaths due to Covid-19 are lower among young people, including women and children, but higher among the elderly and people with chronic diseases. The pandemic appears to have lower local transmission and mortality rates in Africa, the region with the youngest median age of the population and where the virus arrived last. While special efforts should be made to protect the elderly and infirm from infection, preventive measures among women (especially pregnant women) must provide access to emergency care to prevent the maternal mortality caused by Covid-19.

Similar to the crisis of the late 2000s, the current crisis will have an impact on international relations. The structural changes already seen in the globalisation process can be expected to accelerate. In general, the world needs serious investment in research and development to understand current epidemics and to prepare for possible future ones. We need to prepare our health care infrastructure, develop new diagnostic and therapeutic solutions, invest in broad-spectrum vaccines and antivirals, and fund research infrastructure and pandemic predictability. We need more social science research to help understand the social aspects of the pandemic, to foster engagement and trust in our communities, to improve our education to be more adaptive and to target misinformation. We need each other more than ever with greater compassion, solidarity and collaboration. A global pandemic requires global efforts. There will be future severe pandemics.

## Data Availability

No

